# Structured Diabetes Education: virtual access was as effective as face-to-face access to a structured diabetes education programme (EMPOWER T2n) for people with type 2 diabetes in England

**DOI:** 10.1101/2024.02.27.24303369

**Authors:** Donna Sutton, Richard Palin, Jim Swift, Chris Barker, Claire Pridige, Sudip Ghosh

## Abstract

**Introduction & objectives:** Structured diabetes education (SDE) is an evidence-based intervention for type 2 diabetes. The goal of this study was to compare SDE whether accessed face to face or virtually and determine if any differences existed in key endpoint attainment. This study helps address the absence of evaluations comparing these access modalities.

**Research design and methods:** All data were sourced from English SDE participants themselves, and their General Practices and routinely collected for service evaluation between 2016 and 2023. All data were observational, and all participants accessed usual care. The primary endpoint was the increase in the percentage of patients with glycated haemoglobin (HbA1c) at target [48mmol/mol (International Federation of Clinical Chemistry and Laboratory Medicine) / 6.5% (National Glycohemoglobin Standardization Program)] in virtually accessed SDE participants (V-SDE) versus face-to-face accessed SDE participants (F2F-SDE) on unchanged medicines for glycaemia. All data were non-normally distributed. Wilcoxon signed rank tests were used to analyse paired data, Mann-Whitney U-tests used for independent data and Chi-square tests used for observed versus expected data.

**Results:** The 3,493 SDE participants with pre and post HbA1c data had a 10.2mmol/mol (16.4%) reduction in HbA1c, 389 days post their pre-SDE HbA1c measure. In the 2,334 (66.8%) participants who remained on the same medicines regime, the mean reduction in HbA1c was 9.1mmol/mol (15.2%), (p<0.001). All 617 V-SDE participants had a mean reduction in HbA1c of 13.6mmol/mol (20.9%) vs. 9.5mmol/mol (15.3%) in all 2,876 F2F-SDE participants, (p<0.001). The V-SDE on unchanged medicines had superior reductions in HbA1c to F2F-SDE (11.6 [n=404] vs. 8.6mmol/mol [n=1930], p=0.019), respectively. The overall increase in medicines for glycaemia was +12.45% F2F-SDE versus +4.21% V-SDE, (p<0.001).

The primary endpoint was the increase in the percentage of patients with HbA1c to target in V-SDE versus F2F-SDE in patients with unchanged medicines for glycaemia. Previous database analyses found a 30% increase in F2F-SDE patients at target who were on the same medicines regime. A non-inferiority limit was set at 10% for V-SDE versus F2F-SDE and required 360 patients per arm. The primary endpoint was attained with 52.2% of V-SDEs at target (+33.7%), versus the F2F-SDE gain of 29.6%. VSDE was not superior to F2F-SDE (p=0.16). Blood pressure, total cholesterol and weight were improved (all endpoints, p<0.001) with no differences between the interventions. Medicines use was unrecorded for these health endpoints.

**Conclusions:** V-SDE met its non-inferiority goal, which was set in a population in which fewer V-SDE patients required increased medicines for glycaemia. These endpoints were subject to the limitations of unlinked, and routinely collected observational data.

## Background & Introduction

Structured diabetes education (SDE) has sought to enhance participants’ knowledge, skills and confidence to improve the self-management of their diabetes with the aim of improving patients’ quality of life and glycaemia.

The United Kingdom’s National Institute for Health and Care Excellence (NICE) recommended, “Adults with type 2 diabetes are offered a structured education programme at diagnosis”^1^.

EMPOWER T2n both face-to-face (F2F-SDE) and virtual (V-SDE) were externally accredited by QISMET for newly diagnosed patients with type 2 diabetes^2^.

Both interventions had the same content, materials and educators. They were both conducted in a group setting with an average of 12 attendees per session. The only differences were the setting, and access medium.

The study sought to establish if there were any meaningful differences between the two different delivery methodologies of EMPOWER T2n on clinical endpoints and in particular the attainment of 48mmol/mol (6.5%) as the HBA1c target, and additionally patient quality of life and preferences.

This evaluation compares the results of each against the other and both when aggregated against baseline measures. This analysis is a service evaluation of routinely collected data and all results were observational.

Prior to the onset of the COVID-19 pandemic, SDE in the UK, including EMPOWER T2n was almost exclusively delivered face-to-face. The wholesale transition to digital and or virtual SDE was almost immediate. There was a theoretical underpinning for the potential of virtually delivered SDE^4,5^ with some preliminary results available at the time^6^ and since^7,8^. .The course development process has been described in detail^9^. Understanding the implications of this switch to digitally accessed preventative health care has had on key clinical endpoints versus traditional face-to-face services has not been explored adequately viz the impact on HbA1c, overall health status and what participants observed preferences were since coming out of the restrictions associated with the COVID-19 pandemic in the UK. The National Library of Medicine and Cochrane Library were searched for comparative data and one small study was found that was not adequately powered to detect a difference in HbA1c, but patient preference for digital versus traditional was broadly comparable^10^. Spirit Health delivers both forms of SDE to NHS patients and it and the authors have no vested interest in the outcome of this study.

## Methods

This study is a service evaluation of routinely collected data extracted from UK databases. Data were extracted from 2016 to 2023. All data were observational. Participants were referred into the service by their General Practice and accessed usual care. The findings were subject to potential sources of bias and confounding. The results should be considered in that context. The objective was to determine what if any differential impact that access to the two interventions had on key endpoints.

Spirit Health retains participants’ pseudonymised data. It is held in a set of secured, anonymised or pseudonymised, and both linked and unlinked databases where the data are stored on participants’ clinical endpoints, confidence, HRQoL and feedback linked to their assessment of course quality. The internal Spirit Health information governance requirements meant that participants’ protected characteristics could not be linked to the clinical, confidence, HRQoL endpoints or activity data.

The divide between the protected characteristics and other databases meant that matching of participants endpoints for age, ethnicity, sex etc. could not be conducted.

A data map to illustrate the sources of data and when received is beneath in table 1.

**Table 1:** Data map.

| Database | Protected characteristics | Health endpoints | HRQoL | NHS Friends & Family Test | EMPOWER Course |
| --- | --- | --- | --- | --- | --- |
| Source & number | Patients n=14,702 | General Practices n=3,493 | Patients n=2,273 | Patients n=4,023 | Patients n=14,702 |
| When received 1 | Data at referral / baseline | Electronic referral | Data at baseline | On completion | On completion |
| When received 2 | N/A | Data updated at 6 months | Data updated at 6 months | N/A | N/A |
| When received 3 | N/A | Data updated at 12 months | Data updated at 12 months | N/A | N/A |
| Data Links | Isolated & anonymised data |  |  |  |  |

All data where there were baseline and follow-up results; clinical, HRQoL and course preferences were included unless the clinical markers were implausibly zero, e.g., no living person can have a blood pressure or blood glucose of zero. Zero was retained for health-related quality of life (HRQoL), confidence and scoring of the courses.

Clinical results were dependent upon general practices sharing participants’ data. All diagnoses and treatment choices were made in participants’ general practices or secondary care and all test results were shared from their records in standard UK clinical metrics. All data shared where there were follow up results from participants’ practices were included. Participants could choose to respond with their HRQOL, confidence questionnaires at baselines and after 6 and 12 months or not. As with the clinical metrics, the last result shared was the final one carried forward for analysis. Participants could similarly choose to submit results for their evaluation of course elements and the NHS England Friends and Family test or not post course completion and those elements may have added to the potential for confounding.

Clinical endpoint data was sourced from English General Practices in the following integrated care systems, NHS Hertfordshire and West Essex, NHS Lancashire and South Cumbria, NHS Leicester, Leicestershire and Rutland, NHS Lincolnshire, NHS North East and North Cumbria, NHS Sussex, and NHS West Yorkshire. Participant feedback was sourced directly from course participants themselves and permissions were given to use their anonymised data to improve services and for academic study.

Care was taken with glycated haemoglobin (HbA1c) to establish changes in the utilisation of medicines for glycaemia pre and post access to SDE and analyses were conducted on HbA1c in patients with no change in medicines use to attempt to minimise scope for bias and confounding and was the basis for the pre-specified primary endpoint. Changes in the utilisation of medicines for glycaemia were also analysed.

Data cleaning and analyses were conducted in The Microsoft Corporation’s Excel and statistical tests in Excel and The R Foundation’s R and R Studio.

All the data analysed were non-normally distributed, as determined by the Shapiro Wilk / Francia tests, except blood pressures where paired students’ t-tests were used to evaluate the data. Wilcoxon signed rank tests were used to analyse the other paired data in the baseline (pre-EMPOWER) versus final results (post EMPOWER). The Wilcoxon signed rank tests used a Z distribution, normal approximation and were two tailed. The Mann-Whitney U-test was used to compare independent data, primarily the differences between the interventions, which used a normal approximation with ties correction and were two-tailed. Binary endpoints used the Chi-square test. All alphas were set to 0.05. Mann-Whitney U tests and Wilcoxon signed rank tests do not create standard confidence intervals around a mean, and where conducted no confidence interval is reported.

### Participant demographics

Of the 14,702 participants that completed F2F-SDE or V-SDE EMPOWER, results were recorded for 4,023 (29.7%) participants’ course feedback, clinical results were recorded for 3,493 (23.8%) participants, and HRQoL and confidence for 2,273 (15.5%) participants, see table 1.

The mean age of participants was 63.6. The split was 7,739 (52.7%) male, 6,472 (44.0%) female with 491 (3.3%) with other or blank responses. Ethnicity was self-identified by participants and was split: 21.7% Asian, 2.1% Black, 1.1% Mixed Race, 0.5% Other, 73.7% White. The demographics of completed participants may not relate to those whose results are reported because of the divide between participants protected characteristics database and the other databases. The relative over-representation of Asian (21.7% versus 9.3%^11^) and under-representation of Black (2.1% versus 2.5%^11^) and White (73.7% versus 81.7%^11^) participants versus the English population reflected the background demographics of the areas where EMPOWER was most highly accessed. No details on socio-economic status were available for evaluation.

### Statistical testing -clinical endpoints

#### HbA1c and medicines utilisation

The following analyses were conducted; HbA1c total population baseline versus final result, HbA1c F2F-SDE baseline versus final result, HbA1c V-SDE baseline versus final result, HbA1c in participants whose medicines regime for glycaemia was unaltered both F2F-SDE and V-SDE baselines versus final results. The percentage with an HbA1c at target at baseline versus final in both F2F-SDE and V-SDE . The differences in all the HbA1c pre / post tests were also compared between the two groups. All HbA1c measures were reported in mmol/mol. Medicines utilisation and the extent to which medicines increased, remained the same or decreased in the F2F-SDE and V-SDE groups were compared.

To understand if V-SDE was non-inferior to F2F-SDE a treatment to target non-inferiority power sample was calculated. It was assumed that the gain in treatment to target would be around 30% of participants from historic analyses of F2F-SDE HbA1c results. It was accepted that some participants could only access the V-SDE intervention because of limitations in access, e.g., working, or caring responsibilities or difficulties with accessing services related to transport, rurality or disabilities. What would participants be willing to trade off in relation to blood glucose effects? The authors arrived at a 10% reduction for the non-inferiority margin as being acceptable. 20% of potential participants were estimated by call handlers to have rejected the F2F-SDE course for reasons associated with travel or other commitments, e.g., caring or working. It was assumed that both services would be equivalent in the improvement in treatment to target at 30%, and a 10% non-inferiority limit was set. The study required 360 participants in each arm to enable 90% certainty that the upper limit of a one-sided 95% confidence interval excluded a difference in favour of F2F-SDE of more than 10%. The data analysis was not run until the results of over 360 participants in the V-SDE arm who had not had a change in their medicines regime and had baseline and final results were included in the health endpoints database.

#### Other health related endpoints

Systolic and diastolic blood pressures, weight, total cholesterol, and smoking status were compared pre and post and F2F-SDE versus V-SDE. No attempt was made to establish if there were changes in the medicines regime or not for these endpoints nor access to bariatric surgery or other interventions that may have altered these results.

### Statistical testing -other endpoints

For health related quality of life (HRQOL), The EuroQol Group’s EQ tool was used to establish if there were any differences in the two elements of the test; the visual analogue scale [EQ-VAS] and the preference based 5 domain index value [EQ-5D 5L]. Both elements are reported separately. The EQ VAS takes values between 100 (best imaginable health) and 0 (worst imaginable health), on which patients provide a global assessment of their self-assessed health status. EQ-5D 5L index value is a preference-based, i.e. health states were weighted by their perceived disutility and based upon a representative sample of the UK population’s preferences, measure of utility across five domains: mobility, self-care, usual activities, pain / discomfort, and anxiety / depression. The questions were answered by participants at baseline and later at 6 and / or 12 months with the final answer-set used as the comparator with the pre-course baseline.

Participant feedback on elements of the course; the programme, the materials, the educators, and the venues / setting and were scored between 0, the lowest score and 10, the highest score and were compared F2F-SDE versus V-SDE scores.

The NHS England Friends and Family Test was used to determine how valuable patients’ considered the course. It is a structured test that enables patients to evaluate the quality of services provided in a simple, easily understood method. The wording changed, but participants were asked the original question to maintain consistency over time. Participants were asked immediately after completing the course, “We would like you to think about your experience of the EMPOWER course. How likely are you to recommend it to friends and family if they needed similar treatment?” Participants could answer “extremely likely”; “likely”; “neither likely or unlikely”; “unlikely”; “extremely unlikely”; or “don’t know”. The Friends and Family test responses were transformed to numeric scores between 1 extremely likely to 5 extremely unlikely and 6 don’t know and compared between the F2F-SDE and V-SDE respondents. These questions were completed by participants immediately after completion of the intervention.

Activity levels, referral and attendance rates were compared to the English SDE averages for the last available published data in September 2023.

This study followed the RECORD reporting guidelines framework^12^, see appendix 1.

### Bias and Confounding

The inability to link the personal characteristics to the endpoints and match participants meant there was significant scope for confounding in reporting the findings. Different access routes will have excluded different potential participants. The vast majority of participants had no choice over whether to access F2F-SDE or V-SDE, as it was only ever optional after the COVID-19 pandemic had run its course. Before the pandemic, all courses were F2F-SDE and during it all were V-SDE. These sources cannot be meaningfully addressed. Finally, there were clinical results for only 23.8% of all SDE participants. GP Practices could choose to submit data on participants health endpoints or not.

Focusing on participants who had unchanged medicines regimes for the HbA1c results with adequate statistical power to demonstrate non-inferiority helped to minimise additional potential risks of bias or confounding. All courses were delivered whether V-SDE or F2F-SDE by the same team of educators with the same course content and the same materials, albeit in different media and settings.

## Results

There were 3,714 patients with records in the clinical database for HbA1c, they were split 3,075 (82.8%) who accessed F2F-SDE and 639 (17.2%) who accessed V-SDE. Of these 2,876 (93.5%) and 617 (96.6%) had valid pre-post data and were available for analysis.

### Clinical Endpoints – HbA1c & utilisation of medicines for glycaemia

The primary clinical goal of SDE is to reduce blood glucose in participants. And the change in HbA1c can be seen in table 2 beneath for both F2F-SDE and V-SDE participants. The 3,493 EMPOWER participants with pre and post HbA1c data had a 10.2mmol/mol (16.4%) reduction in HbA1c, 389 days post their last HbA1c measure prior to accessing SDE.

**Table 2:** All participants mean results cut by method of access.

| Access method | Baseline HbA1c mean & (95% CI) | Final HbA1c mean & (95% CI) | Difference mean & (95% CI) | Days between measures | No. of participants | p Value* |
| --- | --- | --- | --- | --- | --- | --- |
| F2F-SDE | 61.7 (61.0-62.4) | 52.2 (51.7-52.7) | 9.5 (8.7-10.2) | 406.8 | 2876 | p<0.001 |
| V-SDE | 65.2 (63.6-66.9) | 51.6 (50.6-52.7) | 13.6 (11.9-15.3) | 306.7 | 617 | p<0.001 |

\* Wilcoxon Signed-Rank-test, using Z distribution and normal approximation (two-tailed)

It is evident from table 2 that HbA1c was reduced in participants who accessed both interventions. The difference in baseline HbA1c was statistically different with the V-SDE participants having a statistically higher baseline HbA1c (p<0.001). Participants’ results who had an unchanged medicines regime pre-post and F2F-SDE vs. V-SDE are shown in table 3 beneath.

**Table 3:** HbA1c in participants with unchanged medicines regime.

| Access method and HbA1c changes | Baseline HbA1c mean & (95% CI) | Final HbA1c mean & (95% CI) | Difference mean & (95% CI) | Days between measures | No. of participants | p Value* |
| --- | --- | --- | --- | --- | --- | --- |
| F2F-SDE | 59.8 (59.0-60.6) | 51.2 (50.6-51.7) | 8.6 (7.8-9.4) | 395.4 | 1930 | p<0.001 |
| V-SDE | 61.8 (59.9-63.8) | 50.3 (49.1-51.5) | 11.6 (9.6-13.5) | 311.6 | 404 | p<0.001 |

\* Wilcoxon Signed-Rank test, using Z distribution and normal approximation (two-tailed)

Tables 2 and 3 demonstrate a significant change between the baseline values and final values for both types of interventions and the results were clinically meaningful and statistically significant.

A key underlying component of the primary endpoint was the HbA1c reduction in patients with an unchanged medicines regime. The baseline HbA1cs pre-SDE access were comparable in the unchanged medicines groups (p>0.05), which they were not in the all-participants groups (p<0.05). The results for both comparisons are shown in table 4.

**Table 4:**
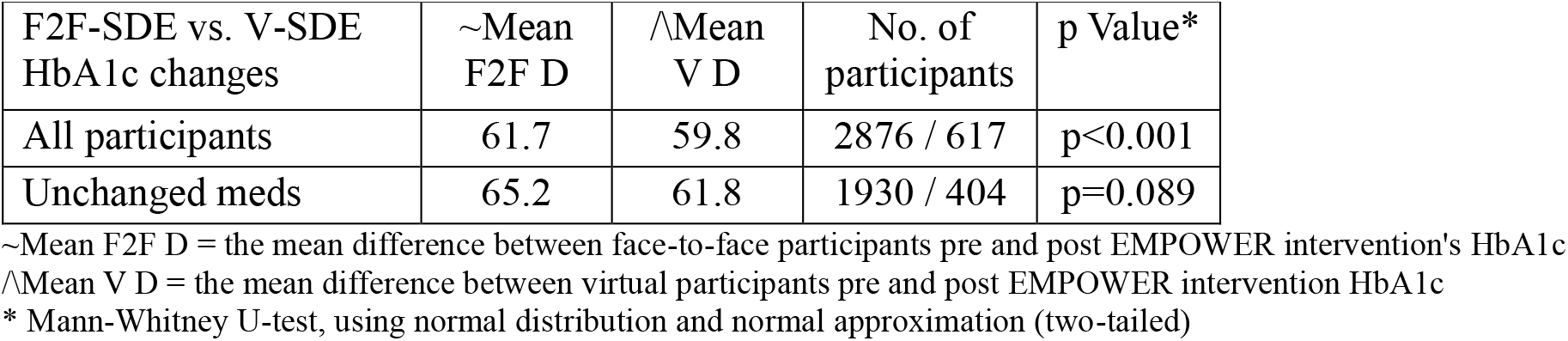
Test to establish if the difference in HbA1c baselines were comparable or not.

### Primary endpoint

A power calculation required 360 participants in the V-SDE group to have baseline and final HbA1c results and 404 were retrieved from the database and available for analysis on the increase in treatment to target (48mmol/mol) / 6.5%). However, while a non-inferiority margin of 10% was set based upon equivalent gains of 30%, the results demonstrated a slightly better effect for the V-SDE intervention (a 33.7% increase from 18.6% at or below their HbA1c target at baseline to 52.2% with their final results) than the traditional F2F-SDE intervention (29.6% increase from 21.8% at baseline to 51.3% at final results). The primary endpoint was achieved. To have achieved the actual results with a 10% non-inferiority limit, the study would have only required 187 patients in the V-SDE group. It is appropriate to conduct both non-inferiority and superiority tests in non-inferiority studies. However, there was no statistical difference between the results, in the superiority analysis (p=0.16), see results beneath in table 5.

**Table 5:** Primary endpoint-test for superiority.

| Categories for primary endpoint | F2F-SDE actual rates | V-SDE observed | V-SDE expected | p value* |
| --- | --- | --- | --- | --- |
| @ target baseline | 0.22 | 75 | 87.9 | 0.16 |
| @ target final result | 0.51 | 211 | 207.4 |  |

\*Chi square test

There was no difference between the baseline HbA1c results (p=0.09), see table 4, in participants with unchanged medicines. Medicines have a greater effect on HbA1c in patients with a higher baseline HbA1c^13,14^. It was hypothesised that structured diabetes education could have a similar effect in patients with higher HbA1c baselines.

In the total population studied, the higher baseline HbA1c levels in V-SDE participants potentially influenced the overall HbA1c change in favour of V-SDE versus F2F-SDE as seen in table 4. In both groups, medicines for glycaemia increased numerically between baseline and final observations. There was no statistical difference in the medicines regime at baseline and final observation for participants in the V-SDE group, which increased overall in 23 patients out of 617 (+3.7%), despite the higher mean baseline HbA1c levels VSDE vs F2F-SDE. In the F2F-SDE group 368 participants (12.8%) had an overall increase in medicines, see table 6 beneath. The difference in overall medication change between baseline and final observation was statistically significantly in favour of V-SDE participants (+4.2%) versus F2F-SDE participants (+12.4%), (p<0.001), as shown in table 6.

**Table 6:**
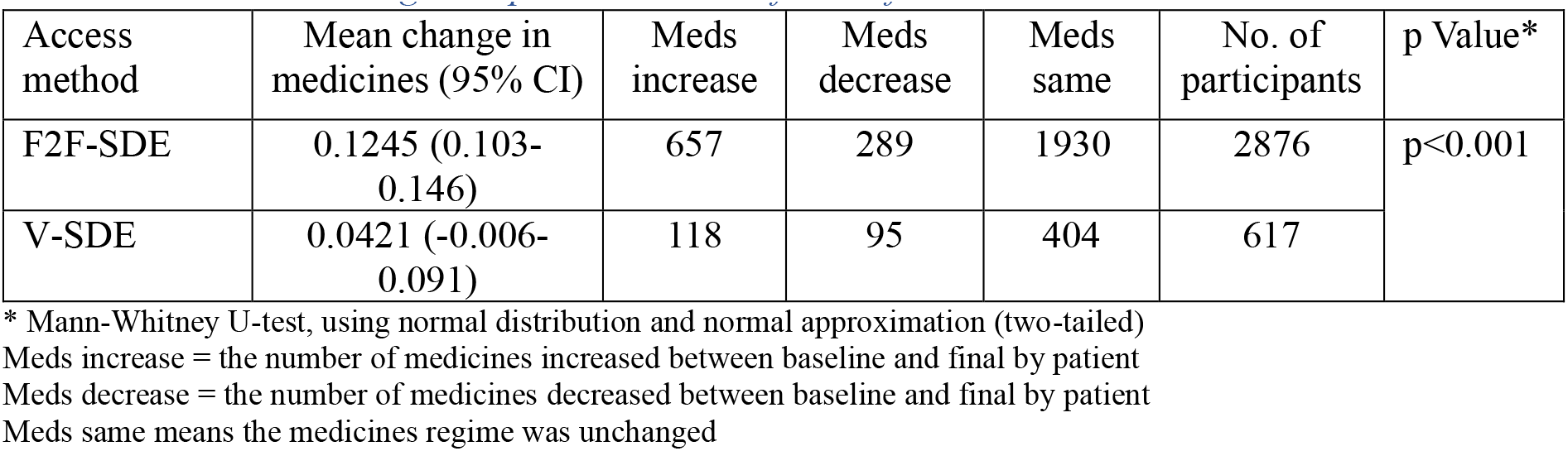
Medication change comparison between face to face & virtual EMPOWER.

| Access method | Mean change in medicines (95% CI) | Meds increase | Meds decrease | Meds same | No. of participants | p Value* |
| --- | --- | --- | --- | --- | --- | --- |
| F2F-SDE | 0.1245 (0.103-0.146) | 657 | 289 | 1930 | 2876 | $p<0.001$ |
| V-SDE | 0.0421 (-0.006-0.091) | 118 | 95 | 404 | 617 |  |

\* Mann-Whitney U-test, using normal distribution and normal approximation (two-tailed)
Meds increase = the number of medicines increased between baseline and final by patient
Meds decrease = the number of medicines decreased between baseline and final by patient
Meds same means the medicines regime was unchanged

### Other clinical endpoints

There was no attempt to measure any change in medicines that could have influenced the outcomes described beneath and as such the influence of medicines or other routinely accessed health interventions for hypertension, lipid metabolism, weight and smoking cessation may have influenced the outcomes.

#### Blood pressure

There were 3,012 valid records (81.1% of all records) in the clinical database with blood pressure results and split 2,599 (84.5%) of all F-SDE records and 513 (80.3%) V-SDE records and had valid pre-post data and were available for analysis.

Systolic and diastolic blood pressures were reduced in both V-SDE and F2F-SDE participants, from 133.3 [95%CI: 132.8-133.8] to 131.0 [95%CI: 130.5-131.5]mmHg^-1^ mean systolic change -2.3 mmHg^-1^, and from 79.7 [95%CI: 79.4-80.1] to 77.8 [95%CI: 77.5-78.2]mmHg^-1^ mean diastolic change -1.9 [95%CI: 1.5-2.3], both p<0.001. There was no difference between the changes in pre / post blood pressures between the F2F-SDE and V-SDE participants for systolic or diastolic blood pressures. A systolic reduction of 2mmHg^-1^ is considered clinically meaningful^15^.

#### Cholesterol

There were 2,034 valid records (54.8% of all records) in the clinical database with both pre and post total cholesterol results and split 1,835 (59.7%) of all F2F-SDE records and 199 (31.1%) V-SDE records available for analysis. There was no difference in the effects on total cholesterol between the F2F-SDE and V-SDE participant results baseline versus final observations between the two groups from 4.6 [95%CI: 4.5-4.6] to 4.2 [95%CI: 4.1-4.2]mmol/L, a mean difference of -0.4 (0.3-0.5). There was a difference in the baseline values (p<0.001) that were higher in the V-SDE group.

#### Smoking

There were 2,112 valid records (57.8% of all records) in the clinical database with both pre and post smoking status results and split 1,777 (59.7%) of all F2F-SDE and 335 (52.4%) V-SDE records available for analysis.

There was no statistical difference in either grouping of participants in final rates of smoking versus baseline observations. Each had positive but small numerical differences. Smoking rates reduced among participants from 9.5% to 9.2% and from 13.1% to 12.2% (both p=0.64) in the F2F-SDE and V-SDE participant groups, respectively. There was a higher baseline of smokers in the V-SDE participants intervention (p=0.043).

#### Weight

There were 1,989 valid records (56.9% of all records) in the clinical database with both pre and post weights and split 1,725 (60.0%) of all F2F-SDE records and 264 (42.8%) V-SDE records and available for analysis.

The baseline weight in both groups was 91.8Kg and reduced to 89.6 and 89.8Kg for F2F-SDE and V-SDE participants respectively; the mean reductions were 2.1 and 2.0Kg (both p<0.001). There was no statistical difference in change between either the groups or their baseline values.

### Health Related Quality of Life

There was no improvement against the preference-based EQ-5D index score in either group of participants. The final versus baseline results were unchanged in both groups pre and post; 0.77 F2F-SDE and 0.74 V-SDE with a difference of-0.001 [95%CI: -0.008-0.006] and 0.0019 [95%CI: -.0283-0.032] and a difference of 351.8 and 278.2 days between measurements in 2,285 and 165 participants with pre and post responses, respectively. Short-run HbA1c changes have demonstrated no improvement in EQ-5D index values in HRQoL^16-18^. EQ-5D index scores are insensitive to small changes in HbA1c but are sensitive to the long run consequences of poor HbA1c control^18-20^. SDE is intended to impact HbA1c and reduce the long run adverse consequences of type 2 diabetes. The changes elicited in patients post SDE have been shown to be highly cost-effective or even cost saving when short-run changes were manifested in long-run outcomes^21-23^.

The EQ-VAS sought participants views on their overall health status. The EQ-VAS baseline values were 68.77 [95%CI: 67.65-69.89] and 57.99 [95%CI: 53.05-62.94] for the F2F-SDE and V-SDE participants with final values of 73.49 [95%CI: 72.59-74.39] and 69.35 [95%CI: 65.66-73.03] changes of 4.62 (6.7%) [95%CI: 3.60-5.84] and 11.35 (19.6%) [95%CI: 6.02-16.69], (both p<0.001) in 2,276 and 165 participants post 351.8 and 278.2 days respectively. The difference between the results favoured the V-SDE participants (p=0.025).

It is worth noting that the EQ-VAS mean baselines were statistically lower in the V-SDE group, (p<0.001). However, the EQ-5D index baseline values were similar in both groups.

### Participant Feedback

In the F2F-SDE NHS Friends and Family Test participants’ results, 99.0% were either extremely likely (84.0%) or likely (15.0%) to recommend the service and 0.2% were either unlikely (0.0%) or extremely unlikely (0.2%) to recommend the service.

98.1% of V-SDE participants were either extremely likely (83.3%) or likely (14.8%) to recommend the service and 0.8% were either unlikely (0.3%) or extremely unlikely (0.5%) to recommend the service. The results are shown in table 7 beneath. The results for the V-SDE participants were statistically similar, mean F2F-SDE 1.18 [95%CI: 1.16-1.20] versus V-SDE 1.22 [95%CI: 1.17-1.26], (p=0.60).

**Table 7:** NHS Friends & Family Test Results.

| Friends & Family Test Results | Extremely Likely | Likely | Neither likely or unlikely | Unlikely | Extremely Unlikely | Don't know |
| --- | --- | --- | --- | --- | --- | --- |
| F2F-SDE (n) | 2841 | 507 | 22 | 1 | 6 | 6 |
| V-SDE (n) | 533 | 95 | 3 | 2 | 3 | 4 |
| F2F-SDE (%) | 84.0% | 15.0% | 0.7% | 0.0% | 0.2% | 0.2% |
| V-SDE (%) | 83.3% | 14.8% | 0.5% | 0.3% | 0.5% | 0.6% |

Participants were also asked to rate four elements of the course they attended: the programme, the materials, the educator, and the venue / setting. The F2F-SDE participants allocated higher scores to the programme (9.4 [95%CI: 9.4-9.5] vs. 9.3 [95%CI: 9.2-9.3], p<0.001) and materials (9.4 [95%CI: 9.3-9.4] vs 9.1 [95%CI: 9.0-9.2], p<0.001), a lower score to the venue / setting (8.9 [95%CI: 8.9-9.0] vs. 9.1 [95%CI: 9.0-9.3], p=0.013) and a similar score to the educator (9.6 [95%CI: 9.6-9.7] vs. 9.7 [95%CI: 9.6-9.7], p=0.876) than the V-SDE participants. Three of four areas had statistical differences, but the actual differences were small and of questionable importance.

### Activity

In 2019 and 2020, 147,810 (69%) and 113,955 (65%) of newly diagnosed patients with type 2 diabetes in England were referred to SDE^24^. Of those 27,090 (18.3%) and 13,285 (11.7%) attended SDE within 12 months^24^. 2019 was not impacted by COVID-19 in the UK, but 2020 was. In 2019 and 2020 there were 5,263 and 3,585 referrals to EMPOWER and 2,575 (48.9%) and 1,604 (44.7%) attended, which equated to a relative performance of 267.0% and 383.8% in converting referrals into attendances, (both p<0.001, Chi square test).

Mechanisms for encouraging patients to attend EMPOWER courses used by Spirit Health are noted beneath.

- Professional call handlers, not clinicians as is common in the NHS and whose training, skills and expertise lie elsewhere, engage with patients using a mix of phone, SMS texting, email and direct mail to encourage patients to attend.
- The call handlers promote the benefits of the course to potential attendees.
- Patients are actively reminded about courses prior to their potential attendance.
- Courses are delivered during the hours of the working week, and also at weekends and in the evenings when it is often more convenient for potential participants.
- For F2F-SDE courses targeted at minority communities, the settings have often been in culturally sympathetic surroundings with a high stress on accessibility (both geographically and on ease of physical access). Courses were delivered in multiple languages.
- Having a virtual intervention that can be accessed by potential participants supported increased access in potentially marginalised groups.

## Discussion

SDE seeks to change behaviour and no behavioural parameters were included in this evaluation only the downstream consequences of potential change; what patients subjectively thought of the courses, their self-assessed health-related quality of life and objective clinical results.

The study was limited because SDE participants were potentially excluded from both F2F-SDE and V-SDE groups during periods when only one access modality was available, which may have added to confounding. There were clinical results for only 23.8% of all SDE participants. GP practices could decide whether or not to submit data for their patients, which meant that over three-quarters of participants had no clinical data reported. The absence of data from such a large cohort of patients was exacerbated by the inability to link key data on protected characteristics, which meant that the groups may have been unbalanced. It is possible that the groups were not matched in important ways that favoured one or other of the SDE courses. There was a substantial potential for differential and non-differential misclassification bias inherent in what data were reported or not and from no data to link the ethnicity and socio-economic status to match participants in each group and no sub-group analysis was possible based on these important characteristics.

Participants in the V-SDE group had higher HbA1c, (importantly, not in the unchanged medicines group) and total cholesterol levels, were more likely to be smokers at baseline, and had lower self-assessed health related quality of life (EQ-VAS) scores at baseline. This suggests a plausible case for the patients not being equally matched with the V-SDE participants being less healthy. However, the overall similarity in course content, the materials, how the courses were conducted and who by and the broad similarity in changes in endpoints in both groups suggested that even if potentially also mismatched in unforeseen ways this may not have adversely influenced the study’s results.

The study achieved its primary endpoint of V-SDE being non-inferior to F2F-SDE in HbA1c reductions to target in patients with an unchanged medicines regime, but not superior. This endpoint reduced any impact of medicines utilisation biasing results. It also favoured F2F-SDE as there was a statistically significantly increased need to add medicines in the population from which this was drawn versus the V-SDE cohort. Against some other metrics V-SDE did demonstrate superiority in HbA1c reductions, but these metrics were not pre-specified, the margins were small and greater numbers accessing V-SDE would have been required to test robustly for superiority. 10% was chosen as the non-inferiority limit. There were reasons associated with equity of access which meant that some participants would be excluded by reasons of geography, access to transport, disabilities, and working or caring responsibilities. The Spirit Health call handlers advised that an estimated 20% of potential participants rejected access to the F2F-SDE version for the aforementioned reasons. Being 10% less effective was considered acceptable. A 10% reduced effect in patients treated to target would be small enough to be more than offset at a population level by the gain in attendance of a sizeable cohort of potential participants.

The aggregated reduction in HbA1c across both groups was 10.2mmol/mol. This reduction was more than twice the minimum clinically important difference designated for new medicines to reduce HbA1c^3^. The HbA1c reductions found in both interventions were what would be considered to be effective.

In regard to endpoints, the V-SDE participants’ health improvements, confidence and self-assessed health status improved by at least as much as the F2F-SDE participants and they scored the course comparably highly.

EMPOWER was able to translate referral into attendance at higher rates than the English national averages; whether F2F-SDE in 2019 or V-SDE in 2020. In the period September 2022 to August 2023 and post the Covid-19 pandemic’s restrictions, V-SDE was observed to be preferred by 71% of participants, which is a strong observed preference and has equity of access implications.

In the age bands 55-64 and over 65, 77% and 69% of people owned a smartphone in 2023^25^ and only 7% of households in the UK did not have access to the internet at home^25^. While access to smartphones and domestic internet access are high in the UK, not everyone did or would choose V-SDE and 29% chose F2F-SDE.

SDE is beneficial^1^ and either extremely cost-effective^21^ or cost-saving^,22,23^ for patients newly diagnosed with type 2 diabetes. 80% of the costs of the management of diabetes were estimated to be associated with managing the downstream costs of its long run complications^26^. SDE can help to reduce the human and financial costs associated with type 2 diabetes complications however it is accessed. EMPOWER T2n may have converted more people from referral to attendance than some other SDE providers, but it still only reached around half of potential participants. Both modalities potentially exclude different sections of society. It is important that many more people diagnosed with type 2 diabetes access SDE.

Given the long-run costs to society of type 2 diabetes, it is suggested that both types of course are made available. Potential participants may choose to forego the undoubted benefits of SDE rather than be compelled to access a course that they are either unable or unwilling to access if both access routes are not made available.

The important conclusions of this study were that virtual EMPOWER T2n was as effective, was rated as highly by patients and when made equally available, it was the observed preference by the majority to face-to-face EMPOWER T2n, but not by all participants.

## Conflict of Interests statement

All authors are employed by Spirit Health Group except for Professor Ghosh who had editorial control, access to and oversight of all data and analyses.

## Ethics

Ethical approval was not required, as this was a service evaluation and was reviewed by De Montfort University Ethics Committee, decision reference FREC HLS 0976/23.

## Funding Statement

All services were delivered to patients in the included NHS Clinical Commissioning Groups and after organisational change by NHS Integrated Care Boards and paid for by the relevant NHS body. No authors were paid over and above their normal salaries for their contributions within their normal roles and no external contractors were paid for additional services. Professor Ghosh was not paid for his time in the oversight of the study design, analyses and its writing.

## This study adds

There is an absence of data comparing digitally / virtually accessed structured diabetes education for people with type 2 diabetes with its traditionally accessed face-to-face comparator on its impact on HbA1c, patient preferences and health related quality of life despite an almost wholesale switch to its virtual access during the Covid-19 pandemic.

## Data availability

Anonymised datasets will be made available to reasonable requests from academic institutions after publication in a peer reviewed journal.

## Appendix 1 RECORD 23 Guidelines

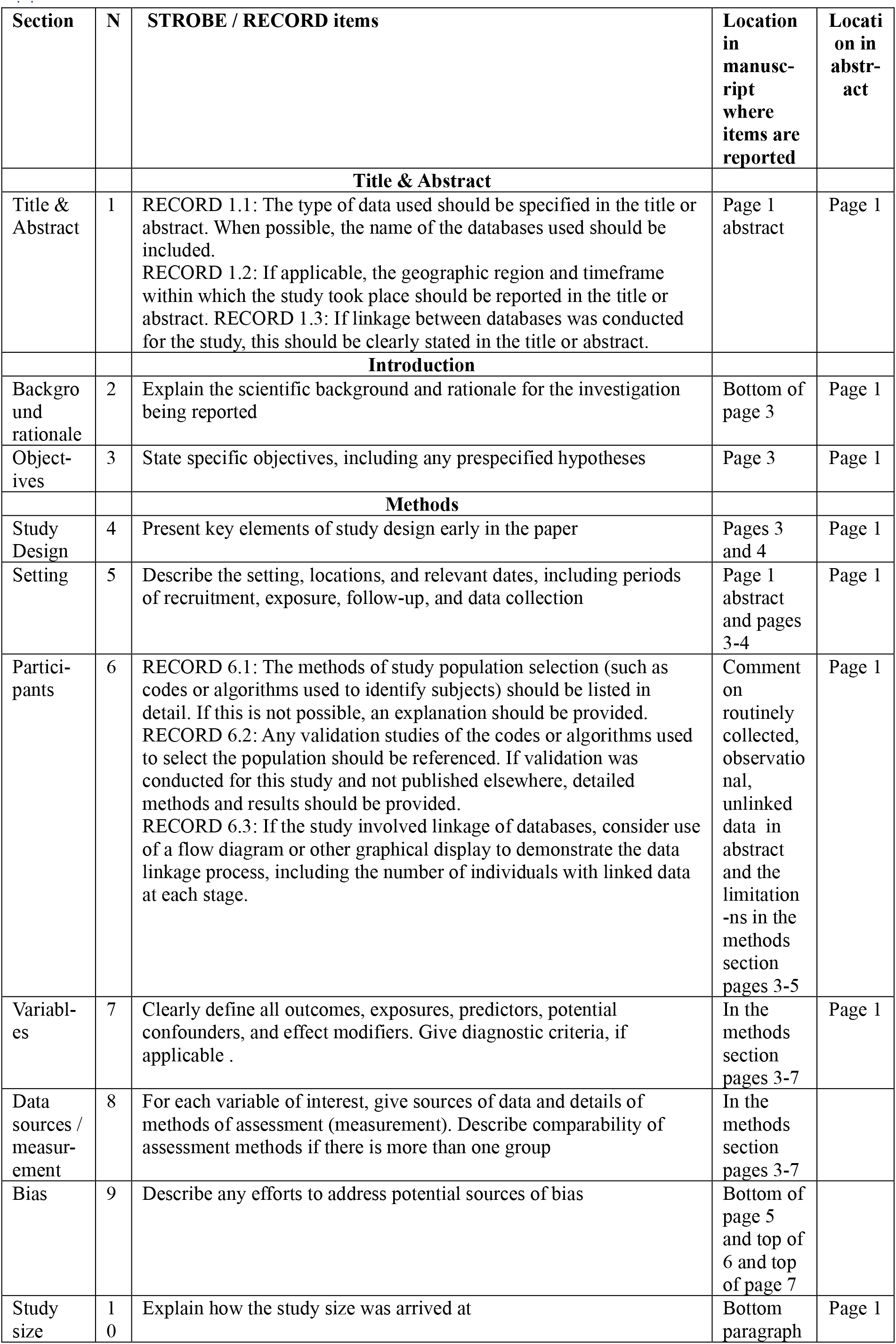
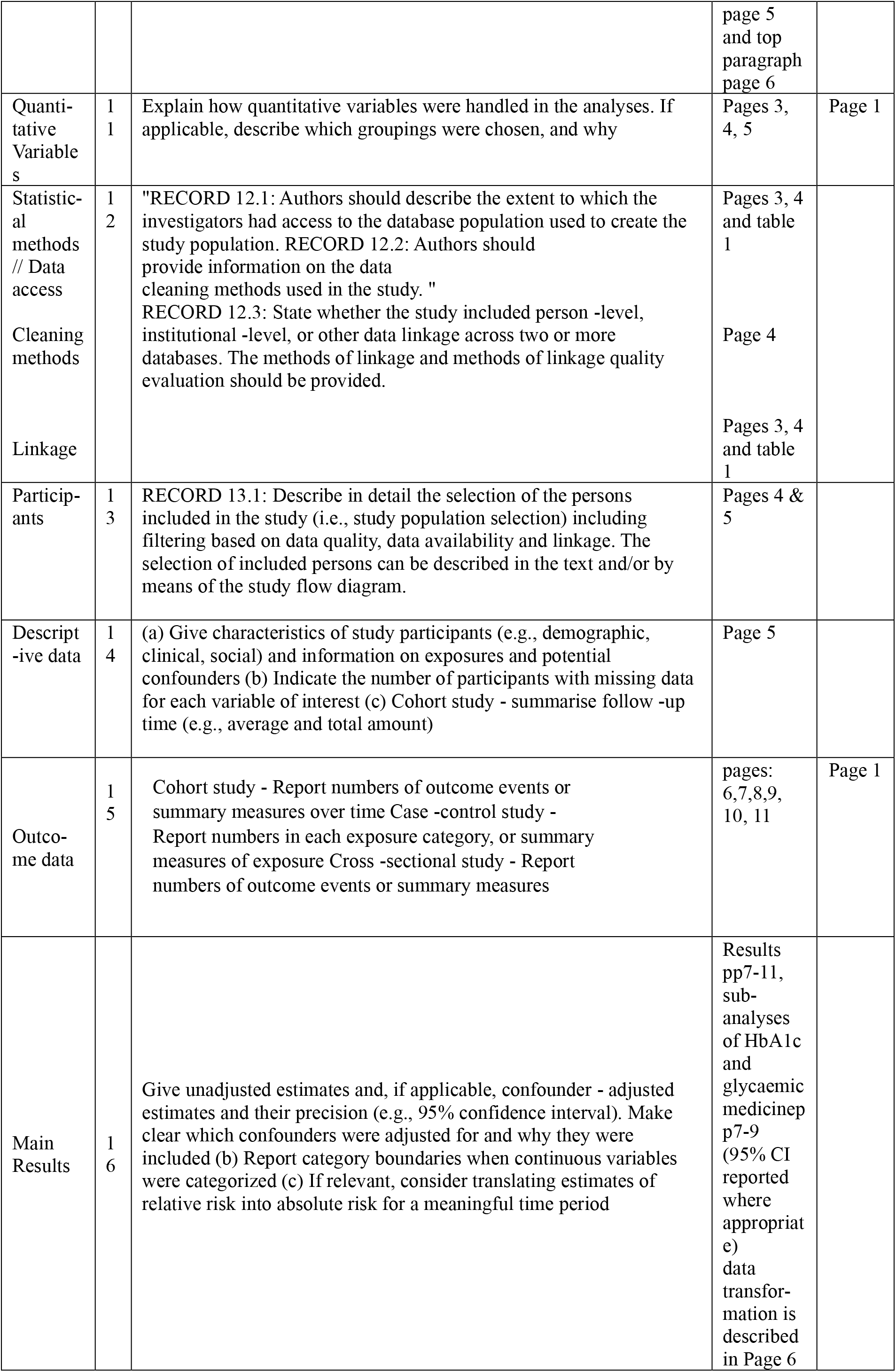
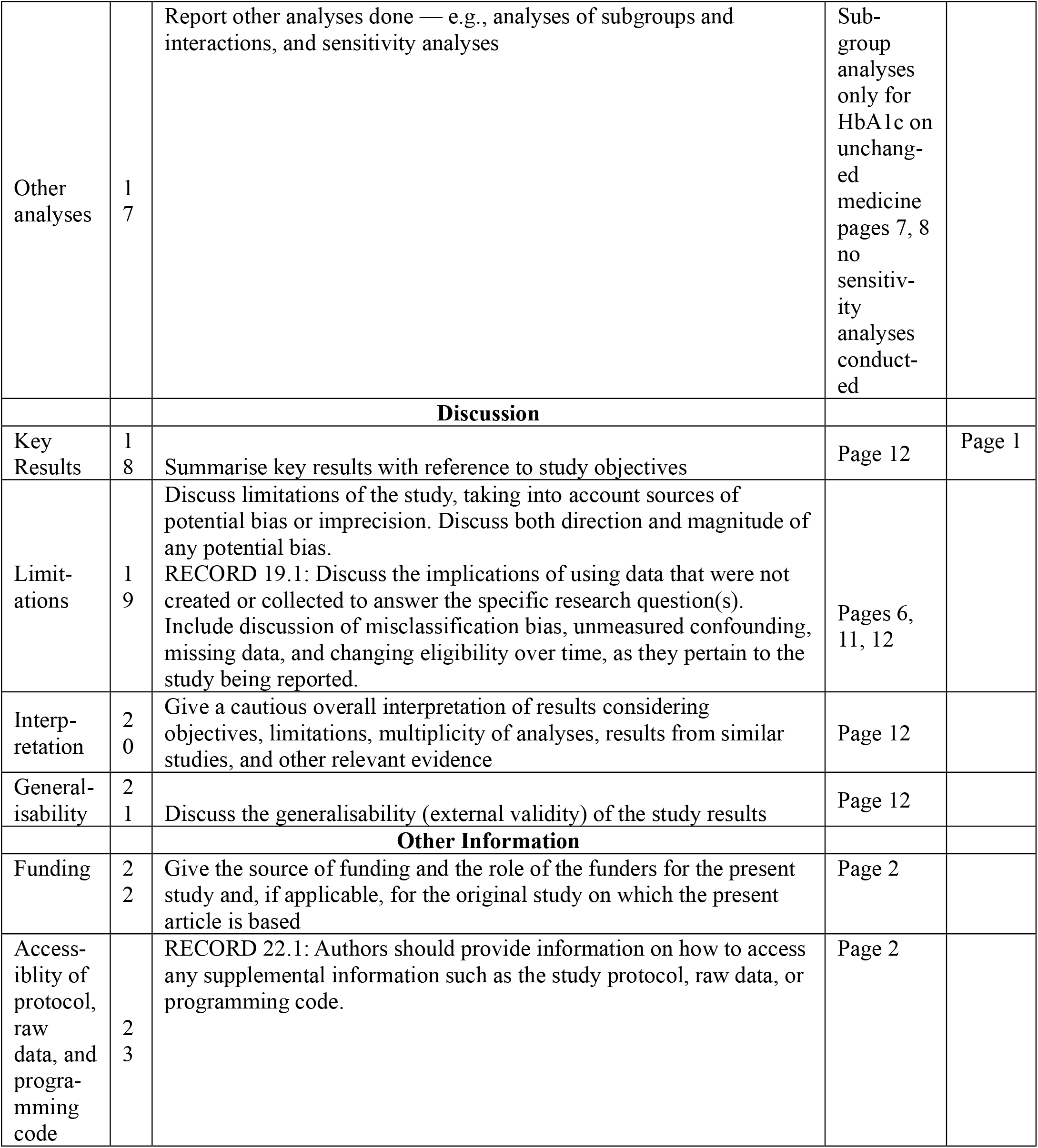

